# An AI-Assisted Comparative GWAS Pipeline Identifies Candidate Sex-Biased Schizophrenia Loci near *CYP26B1* and *EXOC6B*

**DOI:** 10.64898/2026.06.28.26356789

**Authors:** Zhong-shan Cheng

## Abstract

Large genome-wide association studies (GWASs) have generated extensive summary-statistics resources across psychiatric disorders, ancestries, and sex strata. These resources create an opportunity to compare genetic architectures across related datasets, but practical tools for identifying both shared and divergent association signals remain limited. We developed an AI-assisted workflow for local comparative analysis and visualization of multiple psychiatric GWAS summary-statistics datasets. The workflow harmonizes input GWASs, computes pairwise differential association statistics, prioritizes shared loci with concordant evidence across paired datasets, and renders genome-wide and locus-level visualizations with nearby gene context. To improve accessibility and reproducibility, the same analytical workflow can be executed either directly from the command line or through AI-assisted natural-language workflows, while detailed implementation steps remain transparent and locally controlled. We demonstrate the workflow using sex- and ancestry-stratified Psychiatric Genomics Consortium schizophrenia GWAS summary statistics. In the demonstration analysis, the differential workflow highlighted a novel candidate sex-divergent locus at rs185665940 showing protective effect to European females in an intergenic region close to *CYP26B1* and *EXOC6B*, with another independent SNP rs10166057 close to rs185665940 (a risk SNP to schizophrenia and also an brain eQTL of *CYP26B1*) showing female-specific risk association with schizophrenia in both European and Asian female but not male populations. These results show that the workflow can recover biologically credible shared association signals while also identifying candidate subgroup-differential loci for downstream investigation. The pipeline provides a practical bridge between comparative GWAS analysis, publication-style visualization, and AI-assisted reproducible execution under local user control.

## Introduction

Psychiatric disorders are genetically complex, with risk distributed across many loci of small effect [1]. Large consortia, such as Psychiatric Genetics Consortium (PGC), have now generated GWAS summary statistics for multiple disorders and for increasingly refined population strata [2, 3]. These datasets make it possible to ask not only whether a locus is associated with one disorder, but also whether the same locus behaves similarly across related phenotypes, ancestries, and sex strata [1-5]. This comparative perspective is important because psychiatric disorders share substantial genetic architecture while also retaining disorder-specific and subgroup-specific components of risk [1].

Schizophrenia provides a useful example of this challenge. Large GWASs have mapped many schizophrenia-associated loci and implicated neuronal, synaptic, and developmental biology [4]. At the same time, clinical and epidemiological evidence indicates that schizophrenia differs by sex in age at onset, symptom profile, treatment response, and adverse-effect burden [5]. Multi-ancestry resources further show that ancestry-aware analyses can improve interpretation of brain-related genetic signals [6]. These observations motivate workflows that can compare GWAS results across strata such as sex or ancestry while preserving local genomic context by including adjacent genes for detailed evaluation of top associated loci with a specific trait.

Despite the availability of large GWAS resources, practical comparative visualization remains difficult. Widely used tools such as LocusZoom [7], FUMA [8], and the UCSC Genome Browser [9] are powerful for regional visualization, functional annotation, or genome browsing. However, they are not primarily designed to compute pairwise differential effects, prioritize loci that are concordant across paired GWASs, and repeatedly render comparable local views across many user-defined strata. Currently, LocusCompare2 [10] is able to conduct pairwise evaluation of GWASs but lacks the visualization of more than 3 GWASs at once. As a result, investigators often rely on ad hoc scripts, manual file preprocessing, and repeated tool switching.

AI-assisted scientific workflows offer a second opportunity. Large language model agents can help researchers execute repetitive computational tasks, inspect outputs, and rerun analyses with modified parameters [11-14]. However, for biomedical applications, AI assistance is most useful when it is connected to transparent, versioned, and locally reproducible scripts rather than acting as an opaque analytical layer. This is particularly important for genetic workflows, where data provenance, parameter tracking, and reproducibility are essential.

Here we present an AI-assisted workflow for comparative psychiatric GWAS analysis. The workflow was designed to harmonize multiple GWAS summary-statistics datasets, identify shared and differential association signals, and generate publication-ready genome-wide and locus-level visualizations. We provide platform-specific implementation details, installation commands, and Artificial Intelligence (AI)/Model Context Protocol (MCP) registration steps to assist researchers to obtain top common or specific association loci across multiple GWASs. Using sex- and ancestry-stratified PGC schizophrenia GWASs, we show that the workflow can recover established shared loci and identify a candidate sex-differential locus near *EXOC6B*/*CYP26B1*.

## Materials and methods

### Study design and analytical workflow

The workflow consists of three major analytical stages: GWAS harmonization, comparative locus prioritization, and multi-scale visualization (Figure 1). The input is a set of GWAS summary-statistics files representing related phenotypes or strata. The output is a set of ranked shared or differential loci together with genome-wide and locus-specific figures that can be inspected by researchers for further investigation.

**Figure 1.**
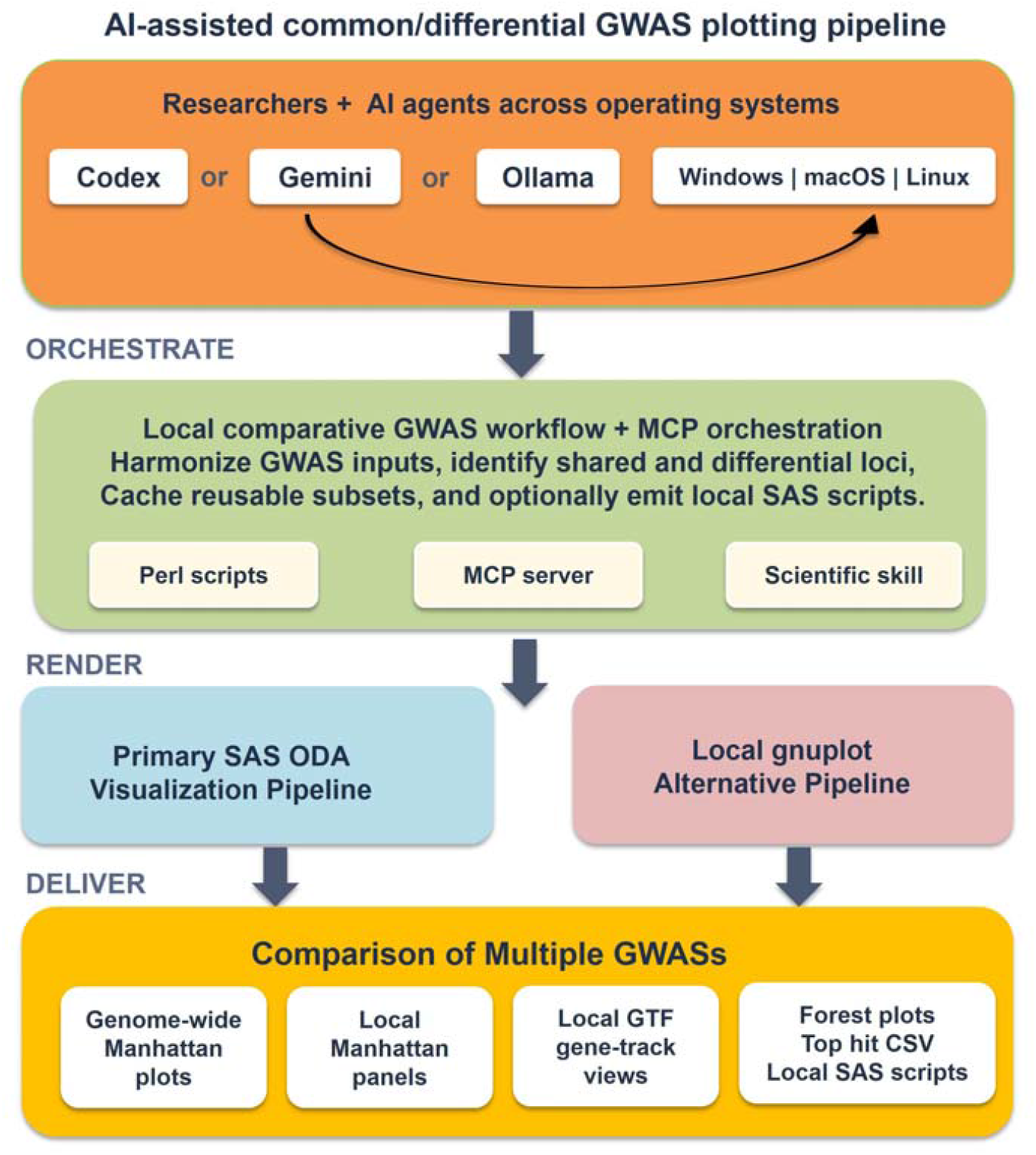
Comparative GWAS analysis with artificial intelligence (AI). Researchers can interact with the workflow through AI agents across different operating systems; the local orchestration layer harmonizes GWAS inputs, identifies shared and differential loci, caches reusable subsets, and can emit local SAS scripts. The rendering layer supports a primary SAS ODA visualization path and a local gnuplot alternative, producing genome-wide Manhattan plots, local Manhattan panels, local GTF gene-track views, forest plots, and tabular or script outputs for reproducible comparative analysis.

The workflow is implemented as a local reproducible pipeline. The computational details, including script names, software dependencies, command-line options, SAS OnDemand for Academics (ODA) [15] execution, local gnuplot [16] rendering, MCP [17] server registration, and cross-platform installation, are provided in the Supplementary Materials.

### Demonstration dataset

The demonstration analysis used sex- and ancestry-stratified schizophrenia GWAS summary statistics [4] from PGC. The bundled example configuration included all-sample female and male GWASs, European female and male GWASs, and East Asian female and male GWASs. These inputs were organized into pooled, European, and East Asian female-versus-male comparisons. The design was intended to demonstrate both differential sex-stratified signals and loci showing concordant association behavior across paired strata.

### GWAS harmonization

Input GWASs were standardized into a common format containing genomic position, allele information, effect estimates, standard errors, and association *P* values. Header aliases and source-specific formats were resolved before merging, allowing downstream analyses to treat all strata consistently. When needed, long-format GWAS tables were converted into compact wide-format tables for plotting and locus prioritization.

### Differential-locus analysis

For paired strata, differential effects were calculated by comparing effect estimates and standard errors between matched GWASs using the Z-score method [18]. Differential beta values, differential standard errors, Z statistics, and standardized differential *P* values were derived for each variant. Loci were ranked according to the configured differential statistic and subjected to distance-based pruning to avoid reporting multiple highly correlated or physically adjacent signals from the same region.

### Shared-locus prioritization

Shared loci were identified using a complementary strategy. A locus was retained when one GWAS track reached a strong association threshold, such as 5e-8, a paired GWAS showed nominal support, i.e. p-value < 0.05, and the paired effects were directionally concordant based on effect size beta or odd ratio. A threshold ladder was used to capture both genome-wide and suggestive signals, followed by distance-based pruning to generate a non-redundant list of representative loci within a 1Mb genomic window. This mode was designed to highlight robust cross-stratum signals rather than subgroup divergence.

### Visualization

The workflow generated three figure families: genome-wide Manhattan plots, locus-specific Manhattan panels, and local gene-track plots. Genome-wide plots provide an overview of association structure across chromosomes and strata. Local plots focus on prioritized loci and display association evidence within a defined genomic window. Gene-track plots add nearby gene and exon context so that candidate loci can be interpreted in relation to local genomic structure. In the main manuscript, we emphasize the genome-wide overview and the most informative local gene-track view for the top differential locus; shared-locus panels are provided in the Supplementary Materials.

### AI-assisted execution of GWAS comparison

The same pipeline can be executed directly from the command line or through AI-assisted workflows (see details in supplementary document). In the AI-assisted mode, an MCP-compatible local server can be exposed for stable analysis actions to AI clients. In addition, a scientific skill is also available for AI clients if user does not want to use the MCP server which requires installation of dependencies. The AI layer helps translate natural-language requests into reproducible local commands, but the scientific logic remains in versioned scripts and configuration files. This design is intended to improve accessibility without sacrificing transparency or local user control.

## Results

### Comparative GWAS workflow output

Our workflow (Figure 1) produced the expected set of comparative outputs: ranked differential loci, ranked shared loci, genome-wide Manhattan plots, local association panels, and local gene-track visualizations. In contrast to a single-GWAS workflow, the output is organized around cross-stratum interpretation. A researcher can first inspect the genome-wide pattern, then focus on differential or shared loci, and finally examine the local genomic context of specific candidate variants.

The genome-wide Manhattan overview from the schizophrenia demonstration analysis shows stacked association and differential-association tracks across autosomes (Figure 2). This plot provides the broad context for prioritizing loci and makes it easier to visually compare signal structure across female, male, ancestry-specific, and standardized differential tracks.

**Figure 2.**
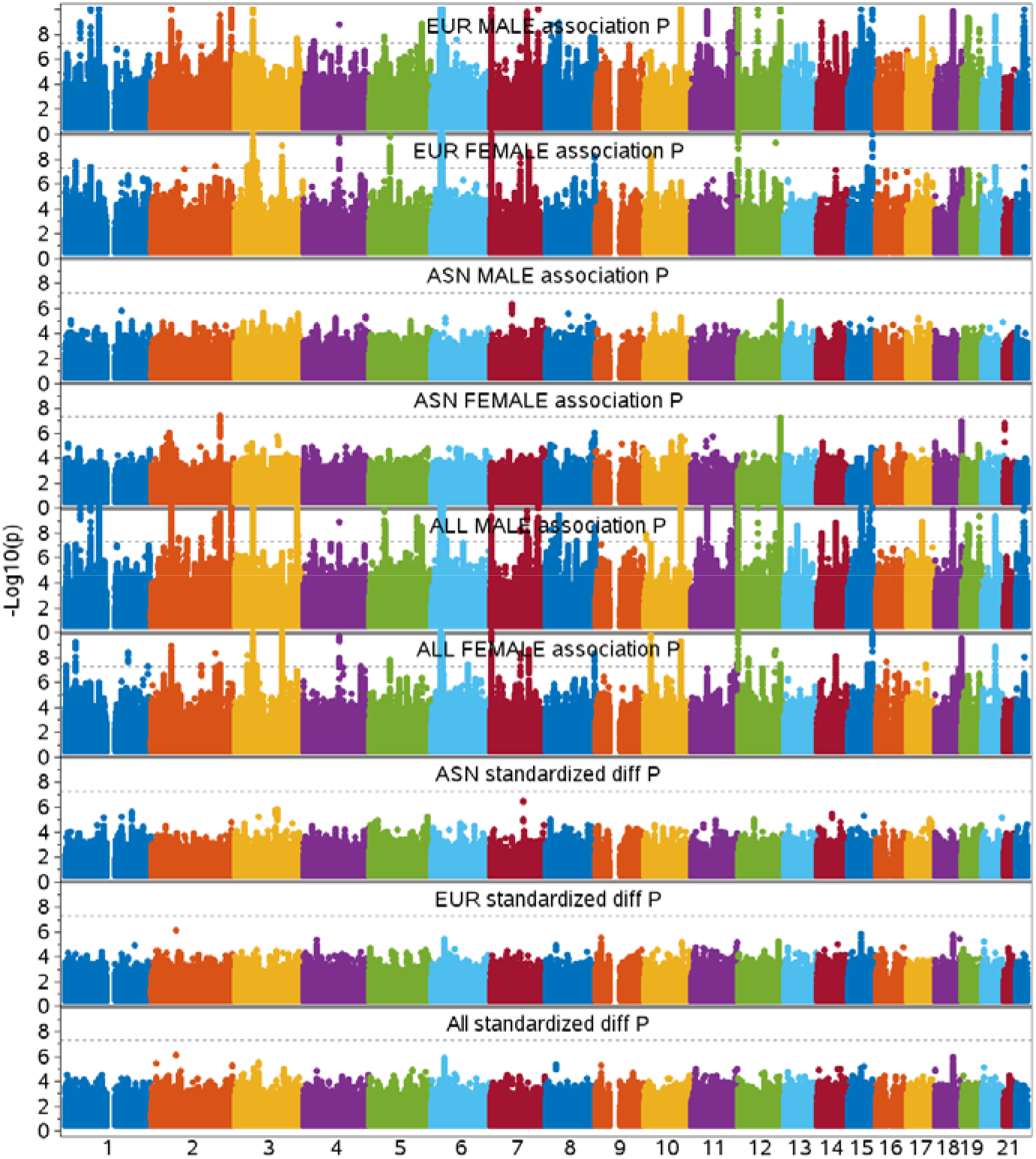
Genome-wide Manhattan plot for the PGC schizophrenia sex- and ancestry-stratified comparison workflow. The stacked layout enables broad comparison of association and differential-association signals across multiple schizophrenia GWAS tracks. The y axis represents -log10(P), with values greater than 10 truncated in the sake of simplicity.

### A candidate sex-differential locus near *EXOC6B*/*CYP26B1*

The differential analysis highlighted rs185665940 as the top candidate locus in the schizophrenia sex-comparison example (Figure 3 and 4). The locus showed a standardized differential *P* value of 7.12e-7, with stronger evidence in the pooled female GWAS (*P* = 2.45e-6) than in the pooled male GWAS (*P* = 0.02), specifically observed in European samples with opposite effect sizes (see the forest plot in Figure 3). According to gnomAD (v4.1.1) [19], rs185665940 tends to be specific to European populations (MAF ∼ 0.01), with much lower MAF observed among other populations, including Asian, African, and Ad Mixed American populations. The plotting workflow labeled the interval to nearby *CYP26B1*, but this label should not be interpreted as definitive causal-gene assignment. Public annotation suggests that the variant is intergenic and physically closer to *CYP26B1* than to the start of *EXOC6B*. In a direct GTEx single-tissue query, we did not identify a public cis-expression quantitative trait locus (cis-eQTL) linking rs185665940 to *EXOC6B* or *CYP26B1*. Nevertheless, Figure 4 also shows distinct association pattern between female and male in combined analysis of all samples, with extra 2 association peaks, represented by rs4852780 (close to *DYSF*) and rs10166057 (close to the promoter region of *CYP26B1*) exclusively observed in female (ALL FEMALE in Figure 4). Furthermore, these two association peaks disappeared in the combined GWAS without stratification by sex. Notably, the association peak close to *CYP26B1*, represented by rs10166057, displays promising association signals (ALL FEMALE *P*=7.81e-05, EUR FEMALE *P*=0.003, ASN FEMALE *P*=0.001; ALL differential *P* between sexes=0.004, EUR differential *P* between sexes=0.05, ASN differential *P* between sexes=0.002), which also shows replicable association with schizophrenia in both EAS- and EUR-females but not males. According to GTEx, rs10166057 is an eQTL of *CYP26B1* in the brain tissue nucleus accumbens (basal ganglia) and an eQTL of the antisense gene ENSG00000289615 of *CYP26B1* in the tissue adipose - visceral (omentum) (supplementary Figure S1). Further sex-biased expression analysis for 3 adjacent genes, including *CYP26B1, EXOC6B*, and *DYSF*, reveals that *CYP26B1* has the largest number of GTEx tissues (n=7, including the brain tissue cerebellar hemisphere) demonstrating nominally differentially expressed *CYP26B1* between sexes. The expression profiles of *CYP26B1* and *EXOC6B* are more similar within each sex group than across sex groups (Figure 5A). Both rs185665940 and rs10166057, as well as rs4852780, were not reported in the original paper as top hits mainly because they demonstrated differential associations with schizophrenia in both sexes and also failed to pass the genome-wide significant threshold in any single schizophrenia GWASs.

**Figure 3.**
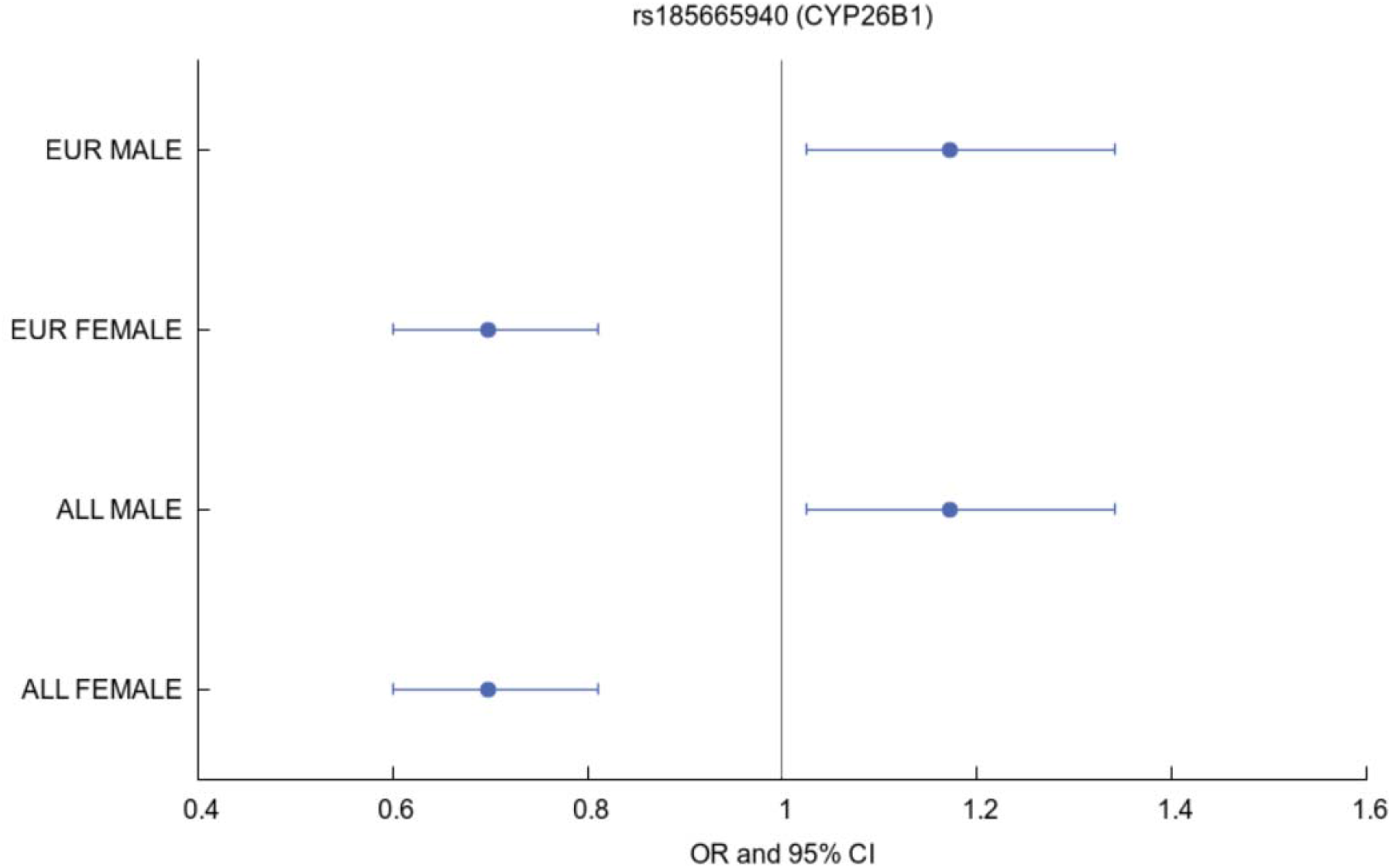
Forest plot for the top differential locus from the PGC schizophrenia sex-comparison example. Due to the minor allele frequency of rs185665940 is only ∼0.01 in European population, the forest plot display almost similar effect sizes between European (EUR) and all population (ALL) combined GWASs of schizophrenia by sex.

**Figure 4.**
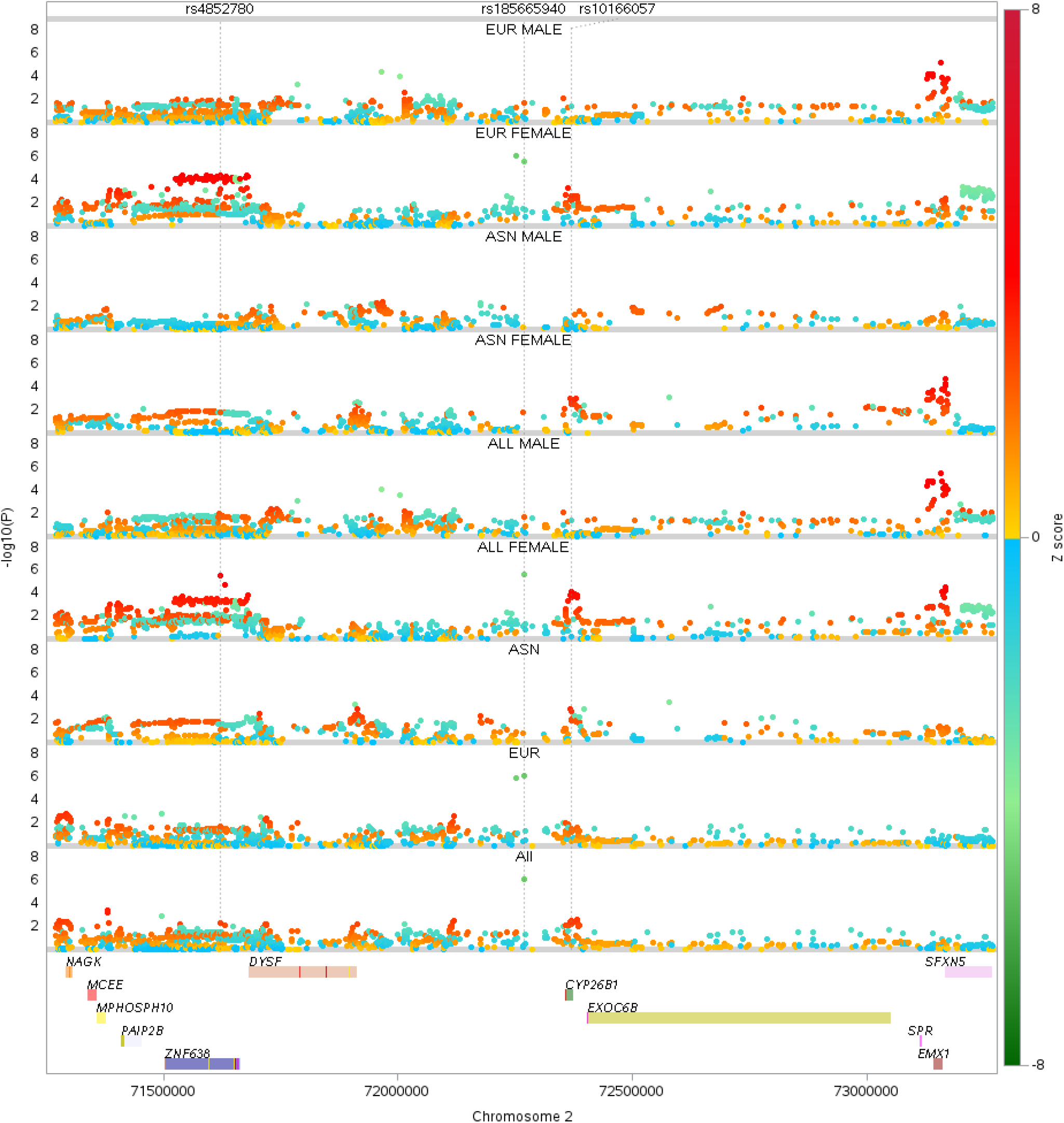
Local GTF gene-track view of rs185665940. The upper tracks display locus-restricted association evidence across schizophrenia strata using -log10(P), whereas the lower track displays nearby gene and exon context. The locus lies in a genomic region close to *CYP26B1* and *EXOC6B*. Note: another SNP rs10166057 representing the peak on the right side of rs185665940 close to *CYP26B1* displays differential association with schizophrenia between sexes in both EUR and ASN populations; there is an another SNP rs4852780 specifically showing stronger association signal in EUR population of female is close to *DYSF*, however, this SNP demonstrates relatively weaker association with schizophrenia in ASN population of female. Data points in the scatterplot are colored according to its corresponding SNP’s association z-score in each GWAS.

**Figure 5.**
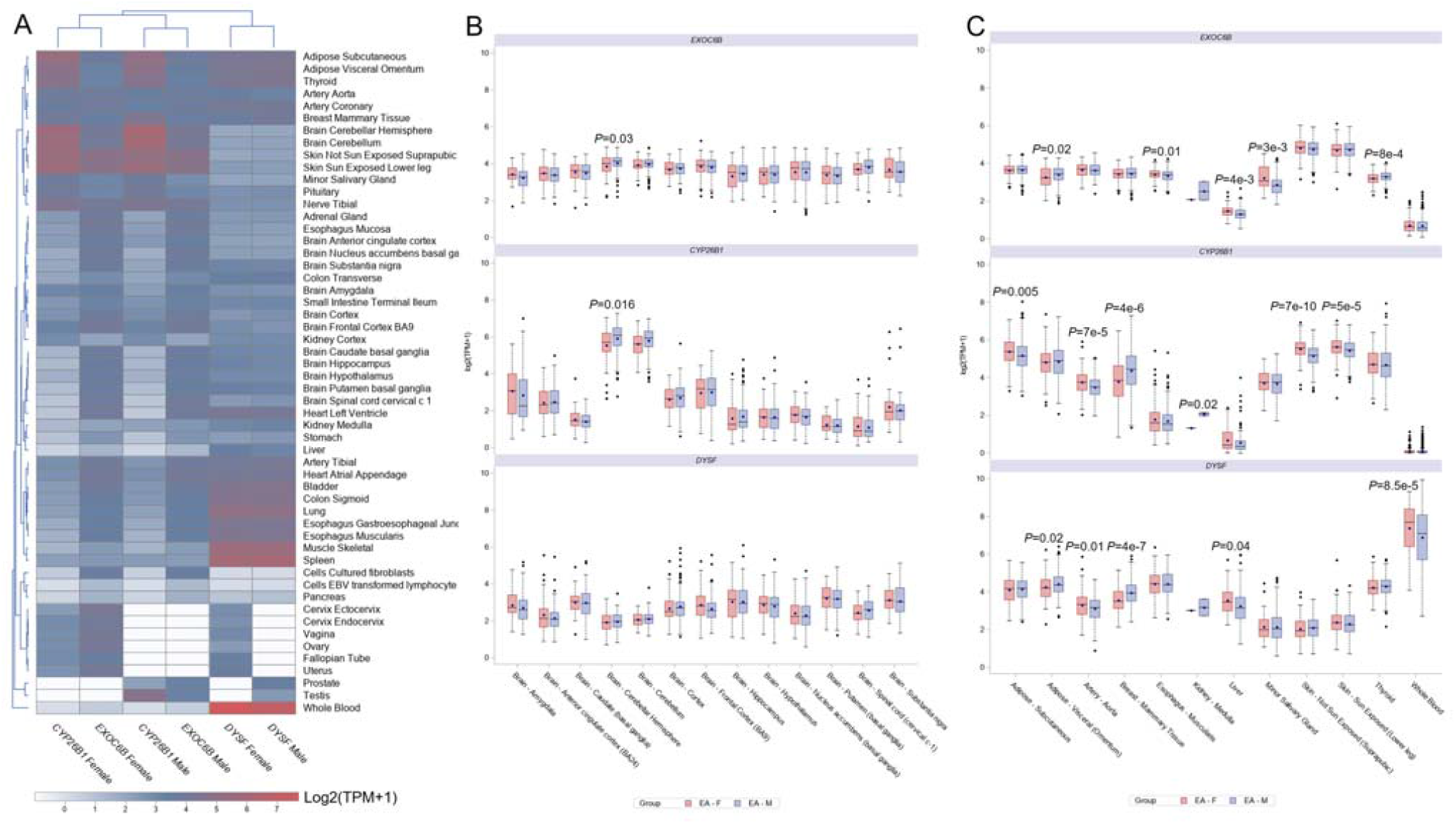
Sex-biased expression of *CYP26B1, EXOC6B*, and *DYSF* among GTEx tissues. (A) Sex-stratified clustering analysis of median expression of 3 adjacent genes of rs185665940 as shown in Figure 4, including *CYP26B1, EXOC6B*, and *DYSF*, among GTEx tissues (n=54), with *CYP26B1* and *EXOC6B* showing more similar expression patterns within the same sex than between sexes. (B-C) Boxplots illustrating the expression of the 3 genes among brain tissues (B) or other non-brain tissues (C) that display at least one nominally significantly differential expression between female and male across GTEx tissues (n=12), and tissues with *P* < 0.05 were explicitly labeled with the differential expression significance. Note: both *CYP26B1* and *EXOC6B* display nominally differential expression between sexes in the brain tissue cerebellar hemisphere.

The biological context of the locus is nevertheless relevant to neuropsychiatric genetics. The genomic region where the top differential SNP rs185665940 resides in has been reported as a schizophrenia risk locus, called Palau 2p13– 14 linkage interval by a previously published linkage study [20]. *CYP26B1*, the most promising candidate gene, encodes a retinoic-acid-metabolizing enzyme expressed in human brain and involved in retinoid homeostasis [20]. *CYP26B1* involves in retinoic-acid signaling during prefrontal cortical development, perturbation of which alters prefrontal patterning, mediodorsal thalamic connectivity, and dendritic spinogenesis in experimental systems [21]. *EXOC6B* encodes a component of the exocyst complex, which participates in vesicle tethering, exocytosis, polarity, migration, and primary ciliogenesis [22, 23]. Disruption or loss of *EXOC6B* has been reported in neurodevelopmentally relevant phenotypes, including developmental delay, epilepsy, intellectual disability, and impaired primary ciliogenesis [22, 24, 25]. In addition, a depolarization-associated circRNA derived from *EXOC6B* has been implicated in activity-linked neural gene-regulatory responses [26]. Compared to the two genes, *DYSF* is mainly relevant to dysferlin-mediated membrane repair, calcium-dependent membrane fusion, skeletal muscle biology, and dysferlinopathy [27-29], suggesting it less likely involved in schizophrenia. Together, these observations support the plausibility of the broader interval while reinforcing the need for cautious target-gene assignment.

### Shared-locus analysis recovers established schizophrenia-associated regions

The shared-association workflow provided a complementary view of the same schizophrenia dataset. Rather than emphasizing sex divergence, this mode retained loci with strong association in at least one track and nominal, directionally concordant evidence in a paired track (Supplementary Figure S2 and S3). Our analysis retained 103 distance-pruned loci spanning 23 chromosomes (see Supplementary Table S1). Representative loci included regions near *VRK2, TEX41, PBRM1, MSL2, PCDH7, CENPE*, and *NMNAT2*. Several of these regions are consistent with the broader schizophrenia GWAS literature, which has implicated synaptic, neuronal, and neurodevelopmental biology [4]. The recovery of known or plausible schizophrenia-associated regions supports the validity of the shared-locus mode.

### Comparative visualization improves interpretability

The main advantage of the workflow is not a single statistical test or a single figure type, but the ability to move rapidly from genome-wide comparison to locus-level interpretation. Genome-wide plots identify regions of interest; ranked tables define differential or shared candidate loci; and local gene-track plots place those variants in genomic context. This integrated flow can help researchers decide which loci should be prioritized for downstream fine-mapping, colocalization, transcriptome-wide association, or experimental validation.

## Discussion

This study presents an AI-assisted workflow for comparative analysis and visualization of psychiatric GWAS summary statistics. The central motivation is that modern psychiatric genetics increasingly involves multiple related GWAS datasets: sex-stratified analyses, ancestry-stratified analyses, disorder subtypes, and related phenotypes. A single-GWAS visualization strategy is often insufficient for these settings because the biological question is comparative: which loci are shared, which are divergent, and how do those signals align with local gene context?

The workflow addresses this need by combining harmonization, shared-locus prioritization, differential-effect analysis, and visualization in one reproducible workflow. Existing tools such as LocusZoom, FUMA, and the UCSC Genome Browser remain essential for post-GWAS interpretation [6-8], but they are not primarily designed to compute pairwise differential signals and repeatedly compare many strata using user-defined rules. Our approach fills this practical gap and is designed to complement rather than replace established resources.

The schizophrenia demonstration illustrates both shared and differential use cases. In shared-association mode, the workflow recovered multiple loci near genes previously implicated or biologically plausible in schizophrenia, including *VRK2, TEX41, PBRM1*, and *MSL2* [4]. In differential mode, the workflow highlighted rs185665940 in an *EXOC6B*/*CYP26B1*-adjacent region. The latter finding should be viewed as hypothesis-generating, because the current analysis does not establish causality, target gene, mechanism, or clinical relevance. Nevertheless, it demonstrates how a comparative workflow can point to loci that deserve additional sex-aware and ancestry-aware follow-up.

The interpretation of rs185665940 also underscores the importance of careful annotation. A nearby-gene label is useful for visualization, but it is not equivalent to causal-gene assignment. The interval includes at least two plausible biological candidates: *EXOC6B*, related to exocyst function and neurodevelopmental phenotypes [22, 24, 25], and *CYP26B1*, related to retinoic acid metabolism in the human brain [30]. Another independent SNP rs10166057 close to rs185665940 displays replicable nominal association with schizophrenia in both EUR and ASN population of female but not male. Also the last SNP demonstrates replicable differential effect sizes between sexes in the two populations. Given that rs10166057 is an eQTL for *CYP26B1* in the brain tissue nucleus accumbens (basal ganglia), future work should be carried out to replicate this locus.

The expanded interpretation of rs185665940 also highlights an important conceptual point for sex- and ancestry-aware psychiatric genetics. In the standardized pooled-sex comparison used here, the effect allele was associated with lower schizophrenia risk in females but higher risk in males, and this sign flip occurred despite very similar retained GWAS-derived MAF estimates in the paired strata (0.014 in females and 0.013 in males). However, the bundled ancestry-stratified output indicates that this opposite-direction signal of rs185665940 is currently European-driven: the retained EUR female and male effect sizes recapitulate the pooled pattern, whereas no corresponding ASN estimate was retained for this SNP. That means the result should be interpreted not simply as a universal sex effect, but more narrowly as a sex-differential signal observed in the European component of the current comparison. Furthermore, the independent SNP rs10166057 tends to be a common genetic factor showing sex-specific risk association with schizophrenia in both EUR and ASN populations of females. A more cautious mechanistic model is therefore that rs185665940 as well as rs10166057 might represent different regulatory haplotypes in the *CYP26B1* region, and that the observed female-protective and male-risk schizophrenia associations arise from sex-dependent regulation of *CYP26B1* with common or unique elements between EUR and ASN populations. One plausible component of that biology is differential control of the *CYP26B1*-retinoic-acid axis within a population-specific regulatory or linkage-disequilibrium background. *CYP26B1* helps spatially restrict retinoic-acid signaling during prefrontal cortical development, and perturbation of this pathway alters prefrontal patterning, mediodorsal thalamic connectivity, and dendritic spinogenesis in experimental systems [21]. A second, non-exclusive possibility is that hormone-sensitive transcription modifies the effect of regulatory variation in this interval. Neuronal Cyp26b1 can be regulated through estrogen-receptor-dependent promoter activity and an antagonistic estrogen-retinoic-acid transcriptional crosstalk [31], which provides a biologically plausible route by which the same local haplotype could be buffered in females yet confer different effects in males. This interpretation is also concordant with broader schizophrenia literature showing that female and male illness biology is not merely quantitatively different but often molecularly distinct, with many illness-related transcriptional and proteomic changes being sex specific [32]. At the same time, these are still mechanistic hypotheses rather than conclusions: the current workflow prioritizes the locus and visualizes its context, but it does not perform formal SNP-by-sex interaction modeling, sex-stratified colocalization, or functional validation.

Several limitations exist in current study. First, the demonstration results are based on repository-bundled example outputs and require independent replication before biological conclusions are drawn. Second, prioritization depends on user-defined thresholds, pruning windows, and annotation choices. Third, the workflow does not currently perform Bayesian fine-mapping, colocalization, expression level association study, or functional enrichment; these are natural downstream extensions. Fourth, although the pipeline supports AI-assisted execution, users still need appropriate knowledge to inspect inputs, evaluate outputs, and avoid over-interpretation.

Despite these limitations, the workflow provides a practical solution for a common analytical problem. By combining comparative GWAS statistics with publication-oriented visualization and AI-assisted reproducible execution, it can support exploratory analyses, manuscript and the prioritization of loci for deeper biological follow-up.

## Conclusion

We developed an AI-assisted workflow for comparative analysis and visualization of psychiatric GWAS datasets or other GWASs. Applied to stratified schizophrenia GWAS summary statistics, the workflow recovered shared loci across strata and highlighted a candidate sex-differential locus near *EXOC6B*/*CYP26B1* with strong signal in European population of female. This workflow provides a practical foundation for future comparative psychiatric GWAS studies and AI-assisted genomic investigations.

## Data Availability

All data produced in the present study are available upon reasonable request to the authors

## Data and code availability

The pipeline scripts and configuration files are available in the GitHub repository (https://github.com/chengzhongshan/MultiGWAS-Explorer).

## Author contributions

ZC created the pipeline and wrote the manuscript.

## Supplementary material

The Supplementary Materials provide implementation details, platform-specific installation guidance, command-line examples, AI/MCP integration steps, backend-specific rendering notes, and supplementary common-association figures and tables.

