## Supplementary material for "An AI-Assisted Comparative GWAS Pipeline Identifies Candidate Sex-Biased Schizophrenia Loci near *CYP26B1* and *EXOC6B*": Revised_Supplementary_Materials_for_V4.docx

### Overview of supplementary material

This supplementary document provides the operational record required to reproduce the analyses, including software components, installation guidance, AI/MCP integration, command examples, backend-specific behavior, and supplementary figures and tables.

The workflow can be used in three modes: direct command-line execution without AI assistance; AI-assisted local execution through an MCP-compatible server; and fully local rendering through the non-SAS gnuplot pipeline when remote SAS ODA execution is not desired.

The pipeline was designed so that AI clients do not contain project-specific scientific logic. Instead, natural-language requests are translated into calls to local, versioned scripts and configuration files. This design supports reproducibility and local user control.

### Major software components

- Local Perl entry scripts: auto_prepare_and_run_diff_gwas.pl and auto_prepare_and_run_diff_gwas_with_gunplot.pl.
- Helper scripts and plotting templates under DiffGWASDeps/.
- Configuration files, including configs/spec_pgc_scz_sex_common_automation.json.
- A Perl-based MCP server, server.pl, for AI-assisted local execution.
- SASPy-mediated access to SAS OnDemand for Academics for the primary publication-style rendering path.
- A local gnuplot/PDL rendering backend for offline rendering and backend comparison.
- Optional local desktop SAS script emission for workstation SAS use.
- Shared dependency manifests, including cpanfile and installations related files.

### Cross-platform installation guidance

The repository includes platform-specific installers for Windows, Ubuntu Linux, and macOS. These scripts provide required Perl and Python environments, install required plotting and compression tools, and reduce the need for users to customize PERL5LIB or manually activate Python environments.

#### Windows portable Cygwin

The recommended Windows entry point is the portable Cygwin bootstrap:

powershell -NoProfile -ExecutionPolicy Bypass -File .\install\install_windows_portable_cygwin.ps1

This wrapper uses MachinaCore/CygwinPortable as an isolated runtime, refreshes required Cygwin packages, writes an isolated fstab, and runs the phase-2 installer inside the portable bash environment. Users already inside a supported Cygwin shell can run bash install/install_cygwin.sh directly.

#### Ubuntu Linux

Run bash install/install_ubuntu.sh. This installer provisions Perl, Python, build tools, bgzip/tabix, SASPy, gnuplot, ImageMagick, and libgd-dev, followed by repo-local Perl and Python dependencies.

#### macOS

Run bash install/install_macos.sh. This installer checks for Xcode Command Line Tools, bootstraps Homebrew when needed, and installs Python, gnuplot, ImageMagick, htslib, GD, pkg-config, curl, and wget before creating repo-local runtime environments.

#### Post-install validation

After installation, users can validate the environment with bash install/check_pipeline_install.sh. The check confirms the availability of gnuplot, ImageMagick, bgzip, tabix, Python imports including saspy and Pillow, Perl modules including GD, Mojolicious, MCP::Server, and Inline::Python, and syntax validity of the main entry scripts and SAS ODA wrappers.

### SASPy and SAS ODA rendering path

The primary visualization path uses SASPy as the bridge between the local computer and SAS OnDemand for Academics. SAS ODA is used mainly as a rendering backend for compact derived GWAS subsets rather than as the engine for full raw GWAS preprocessing. The helper run_sas_codes_or_script_in_ODA.pl supports code submission, SAS file submission, file upload and download, remote file deletion, and remote file metadata queries.

The helper also supports first-run credential bootstrap. If no saved SASPy authinfo entry exists for authkey oda, it can prompt interactively for the SAS ODA account and password, validate the login using proc setinit;run;, and save only successful credentials for later use. Noninteractive login can be supplied through --sas-oda-account and --sas-oda-password. To intentionally refresh a saved login, users can use --prompt-sas-oda-auth.

### Local gnuplot rendering path

The local gnuplot path preserves the same preprocessing and top-hit selection logic but renders figures entirely locally. It is useful when users prefer offline rendering or want to prototype layout changes without relying on SAS ODA availability. The gnuplot backend supports genome-wide Manhattan plots, local Manhattan panels, and local GTF gene-track views, including selected GWAS tracks and explicit inquiry SNPs.

For visual consistency with the main manuscript, chromosome X is removed from final genome-wide figures by default in the bundled gnuplot workflow unless --no-remove-X-chr is supplied. Cached plotting subsets are validated before reuse and regenerated atomically when needed.

### Running the pipeline locally without AI

Example A. Genome-wide, local Manhattan, and local GTF plots in the SAS ODA path:

perl ./auto_prepare_and_run_diff_gwas.pl --spec configs/spec_pgc_scz_sex_common_automation.json --step plot_manhattan --step plot_local_manhattan --step plot_local_gtf --force

Example B. Common-association mode in the SAS ODA path:

perl ./auto_prepare_and_run_diff_gwas.pl --spec configs/spec_pgc_scz_sex_common_automation.json --get-common-associations=1 --step plot_manhattan --step plot_local_manhattan --step plot_local_gtf --force

Example C. One-SNP local GTF test in the SAS ODA path:

perl ./auto_prepare_and_run_diff_gwas.pl --spec configs/spec_pgc_scz_sex_common_automation.json --get-common-associations=1 --target-snps rs4950119 --target-snp-genes rs4950119:ENSG00000294947 --step plot_local_gtf --force

Example D. Single-GWAS plot set in the SAS ODA path:

perl ./auto_prepare_and_run_diff_gwas.pl --spec configs/spec_pgc_scz_sex_common_automation.json --display-gwas ALL_FEMALE --step plot_manhattan --step plot_local_manhattan --step plot_local_gtf --force

Example E. Genome-wide and local views in the local gnuplot path:

perl ./auto_prepare_and_run_diff_gwas_with_gunplot.pl --spec configs/spec_pgc_scz_sex_common_automation.json --target-snps rs185665940, rs4950119 --plots manhattan,local_manhattan,local_gtf

Example F. Emit local runnable SAS scripts for desktop SAS:

perl ./auto_prepare_and_run_diff_gwas.pl --spec configs/spec_pgc_scz_sex_common_automation.json --step plot_manhattan --step plot_local_manhattan --step plot_local_gtf --emit-local-sas-scripts

### Adding the Perl MCP server to AI platforms

The local MCP server can be launched from a compatible shell with:

./server.pl daemon -m production -l http://127.0.0.1:8080

The MCP endpoint is http://127.0.0.1:8080/mcp. For Codex, the endpoint can be registered with codex mcp add perl-bio --url http://127.0.0.1:8080/mcp followed by codex mcp list. The same endpoint can be connected through other MCP-compatible clients, including Gemini- or Ollama-based hosts, such as mcphots, provided the client can access local HTTP MCP tools.

### VS Code-centered AI-assisted workflow

Across Windows, macOS, and Ubuntu Linux, the recommended AI-assisted pattern is to open the repository as the active VS Code workspace, complete the matching platform installer, open an integrated terminal rooted in the repository, start server.pl in one terminal, register the local MCP endpoint with the AI client, and then use natural-language requests to launch the same local pipeline scripts. If the user don’t load MCP, AI agent still can access the pipeline via its scientific skill.

For Windows, the preferred shell is the portable Cygwin environment created by the installer. For macOS and Ubuntu Linux, the native integrated zsh or bash terminal can be used. This editor-centered design keeps the repository, local server, shell environment, and output files synchronized in one workspace.

### Example AI prompts

- Use the PGC schizophrenia sex-comparison spec and generate the genome-wide Manhattan plot, the combined local Manhattan plot, and the local GTF plots for differential-association loci.
- Use the schizophrenia spec, focus on rs4950119, label it with ENSG00000294947, and generate a local GTF plot for that target SNP only.
- Run the gnuplot workflow for rs185665940 and generate the genome-wide Manhattan plot, combined local Manhattan plot, local GTF output, and forest plot.
- Use the schizophrenia spec, display only ALL_FEMALE, and generate genome-wide, local Manhattan, and local GTF outputs.

### Supplementary figures

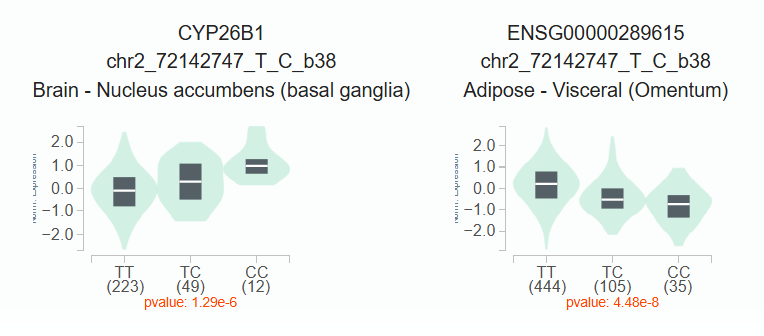

Supplementary Figure S1. rs10166057 is an eQTL of CYP26B1 and its antisense gene ENSG00000289615 in GTEx database. ENSG00000289615 is an lncRNA gene (novel transcript, antisense to CYP26B1).

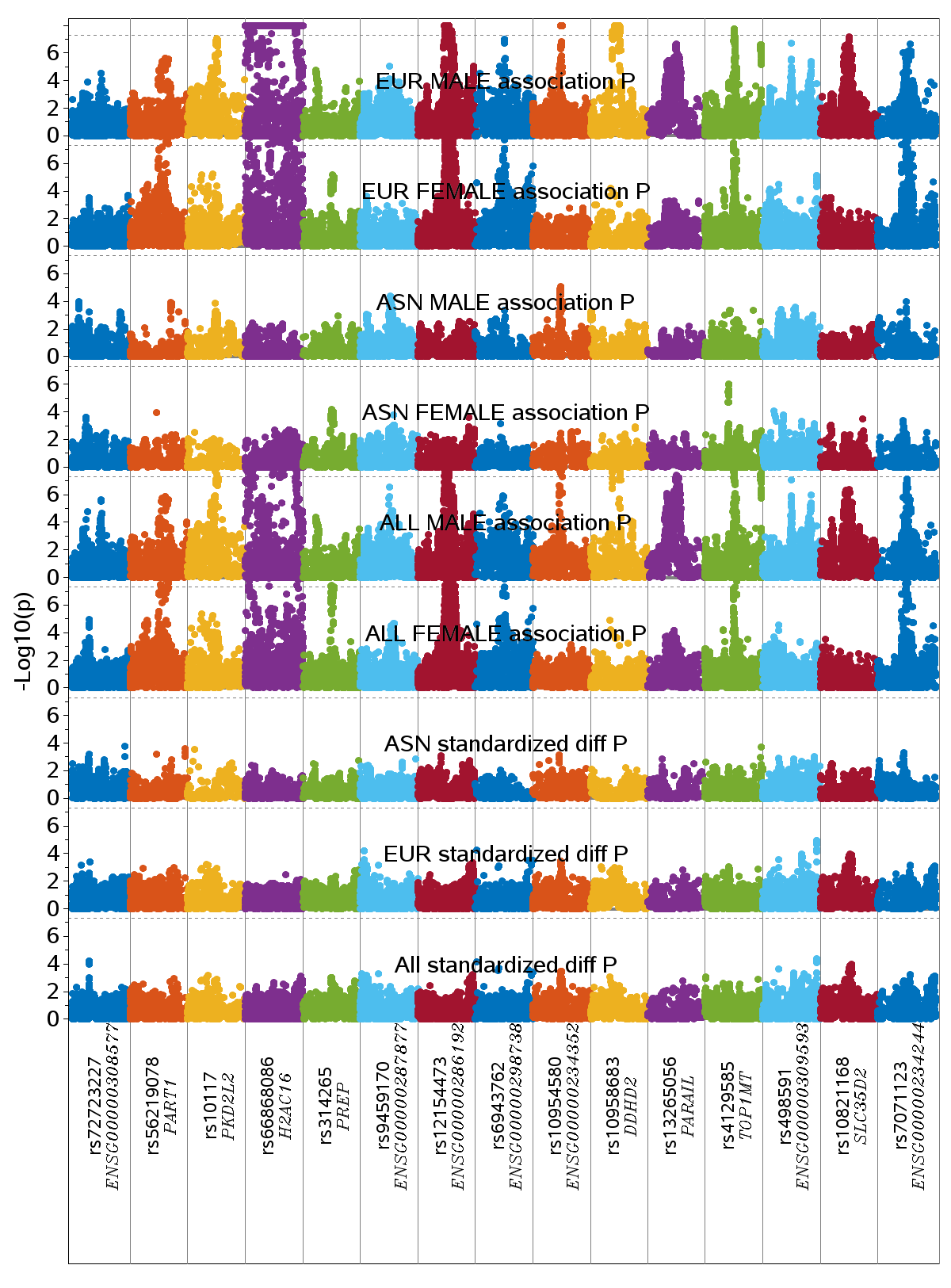

Supplementary Figure S2. Combined local Manhattan panel for representative common top hits from the PGC schizophrenia sex-ancestry workflow. This panel summarizes regional association windows across selected schizophrenia strata and illustrates shared association behavior.

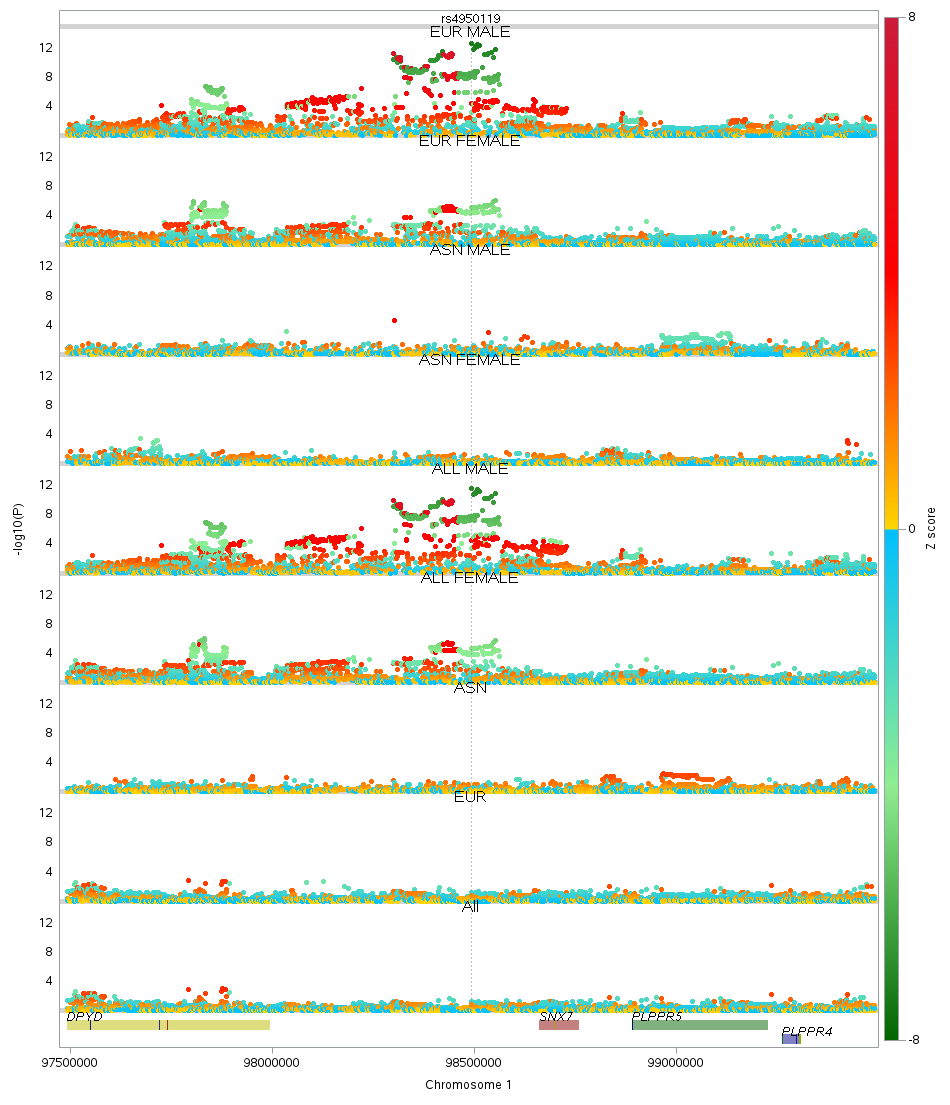

Supplementary Figure S3. Local GTF gene-track view of the representative common-top-hit locus rs4950119. The plot uses the same local GTF rendering framework as the main-text differential example and provides local gene/exon context for a shared-association locus.

### Supplementary tables

Supplementary Table S1. Independent common-association loci from the bundled PGC schizophrenia sex-ancestry example.
